# Combined *MGMT* Expression and MMR Deficiency Underlie Poor Outcomes of Temozolomide in *IDH*-Wildtype Grade 2 Gliomas

**DOI:** 10.64898/2025.12.12.25342173

**Authors:** Ruolan Zhang, Shuai Wu, Lin Chen, Zehui Cao, Erfei Shang, Weicheng Zheng, Chen Luo, Shanyue Sun, Shangchen Xu, Qingwang Chen, Yang Ming, Leming Shi, Yuanting Zheng, Yingchao Liu, Jinsong Wu

## Abstract

**Background:** Isocitrate dehydrogenase wildtype (IDHwt) histologic grade 2 adult diffuse gliomas represent a highly heterogeneous entity, and the effects of postoperative adjuvant temozolomide (TMZ) therapy, as well as the predictive value of chemotherapy-related biomarker O6-methylguanine-DNA methyltransferase promoter (MGMT-p) methylation, remain to be further investigated.

**Methods:** A Discovery dataset, comprising 108 *IDH*wt histologic grade 2 gliomas obtained from three public resources, was constructed to investigate the impact of TMZ on patient survival. Furthermore, an independent Validation dataset, consisting of 123 *IDH*wt grade 2 gliomas, was created to validate the effect of TMZ on patient survival. Kaplan–Meier overall survival (OS) analyses and Cox proportional hazard models were used.

**Results:** Multivariable analysis in the validation dataset demonstrated that temozolomide (TMZ) chemotherapy was an adverse independent prognostic factor for survival in patients with histologic grade 2 *IDH*-wildtype (*IDH*wt) gliomas (HR = 2.19, *P* = 0.033). Subgroup analyses further revealed that the detrimental effect of TMZ was mainly confined to *MGMT*-p unmethylated tumors (Discovery cohort: TMZ vs noTMZ, median overall survival [OS]: 37.0 vs 130.0 months, log-rank *P* = 0.005, HR = 1.89; Validation cohort: TMZ vs noTMZ, median OS: 34.4 vs >120 months, log-rank *P* = 0.004, HR = 3.39). Moreover, in the validation cohort, TMZ therapy remained associated with significantly poorer survival in the NEC (Not Elsewhere Classified) subgroup (TMZ vs noTMZ, median OS: 73 vs >120 months, *P* = 0.017), while in molecular GBMs it did not reach statistical significance but still showed a trend toward worse survival (TMZ vs noTMZ, median OS: 20 vs 30 months, *P* = 0.37). By comparing multi-omics differences between patient groups, we observed that MMR-related genes were specifically downregulated in *IDH*wt grade 2 gliomas at both the single-cell and bulk transcriptomic levels.

**Discussion:** The therapeutic benefit of TMZ in IDHwt histologic grade 2 gliomas appears to be limited, with even a potential adverse impact on survival. Therefore, postoperative use of TMZ as a recommended chemotherapy should be approached with caution in these patients, particularly in cases with unmethylated *MGMT*-p, where alternative treatment strategies are warranted. High *MGMT* expression and specific downregulation of MMR-related genes may represent key factors underlying the limited efficacy of TMZ in *IDH*wt, *MGMT*-p unmethylation grade 2 gliomas.

**Level of evidence:** III.

## Introduction

The optimum chemotherapy regimen for adult patients with isocitrate dehydrogenase wild-type (*IDH*wt) histological grade 2 gliomas remains unclear^1^. This entity encompasses a heterogeneous group of patients characterized by substantial clinical, histopathological, and molecular diversity. In the *2021 WHO Classification of Tumors of the Central Nervous System 5^th^ edition*, *IDH*wt adult diffuse grade 2 glioma is now considered under the category of molecular glioblastoma (a telomerase reverse transcriptase promoter (*TERT*-p) mutation, epidermal growth factor receptor amplification (*EGFR* amp), or combined whole chromosome 7 gain and whole chromosome 10 loss (+7/-10) but without microvascular proliferation or necrosis^2–4^), not elsewhere classified (NEC) or other specified types. Unfortunately, no definitive chemotherapy regimen has been established for either molecular GBMs or NEC patients^5–12^.

Temozolomide (TMZ), as part of the Stupp regimen combining surgical resection, radiotherapy, and adjuvant TMZ, is the mainstay treatment for classic histological grade 4 glioblastomas (GBMs)^13–15^. Based on their molecular resemblance to classic GBMs, molecular GBMs have often been considered for TMZ treatment. However, the benefit of this approach remains uncertain, as these tumors usually have more favorable survival, and supporting evidence for a therapeutic advantage is lacking^16–19^. In contrast, for gliomas categorized as NEC, treatment strategies remain largely undefined, as current guidelines provide no specific therapeutic recommendations^20,21^.

We summarized clinical cohort studies of TMZ treatment from 2003-2023 in **Table S1**. Response rates to TMZ chemotherapy in grade 2 patients fluctuated widely (6%-61%), failing to identify a group suitable for TMZ treatment. The NRG Oncology/Radiation Therapy Oncology Group 0424 (RTOG-0424) trial found that chemotherapy improved 3-year survival in patients with high-risk grade 2 gliomas^22^. In contrast, another phase II clinical trial found no significant overall survival benefit; notably, more than half of patients with grade 2 gliomas with 1p/19q retained and *IDH1*wt in this trial exhibited rapid progression after TMZ treatment^7,8^. Additionally, histological grade 2 samples for *IDH*wt are particularly lacking, and several reports were too small to draw conclusions^20,21^.

In addition to drug effect, the prognostic value of O^6^-methylguanine-DNA methyltransferase promoter (*MGMT*-p) among grade 2 *IDH*wt glioma patients is unclear as well. MGMT is a DNA repair protein that prevents TMZ cytotoxicity by removing the alkyl group from the O6 position of guanine^23^. Methylation of *MGMT*-p represses *MGMT* transcription, reduces *MGMT* protein, and thus serves as a sensitivity marker for TMZ in GBM patients^24–29^. The RTOG-0424 trial reported that *MGMT*-p methylation was an independent prognostic biomarker for low-grade gliomas treated with TMZ and radiotherapy^21^. However, due to the rarity of *IDH*wt low-grade glioma patients in the trial, the effect of *MGMT*-p methylation status on the survival of *IDH*wt grade 2 adult diffuse gliomas was not evaluated^21^. Hence, it is important to determine the prognostic value of *MGMT*-p in a cohort with a larger sample size.

In this retrospective study, we performed hierarchical analysis based on different molecular stratifications to evaluate the effect of TMZ chemotherapy in a large cohort of *IDH*wt histological grade 2 adult diffuse glioma patients, with the aim of identifying subgroups that may truly benefit from TMZ as well as those potentially harmed by its use^30^. Using publicly available datasets and data from two local institutions, we aim to provide insights into prognosis and treatment strategies for this patient group, and to highlight potential factors underlying differences in chemotherapy efficacy. Our research can be considered a first attempt to redefine postoperative treatment for *IDH*wt histological grade 2 adult diffuse gliomas, offering a framework for individualized management and future prospective studies.

## Patients and Methods

### Study populations

The process of case selection is shown in **Figure 1**. Patients diagnosed with “Histologic Grade 2, *IDH*-wildtype” according to histological review were included in the Discovery dataset and Validation dataset. The Discovery dataset (n = 108, **Figure 1**, **Table 1**) was integrated from three public resources: The Cancer Genome Atlas (TCGA, https://www.cancer.gov/about-nci/organization/ccg/research/structural-genomics/tcga), the Chinese Glioma Genome Atlas (CGGA, http://www.cgga.org.cn/), and a Japanese cohort^31^. Patients without available survival, *IDH*, *MGMT*-p, and required clinical (grade, gender, follow-up, and treatment) information were excluded. TMZ was used for patients in the TCGA and CGGA cohorts, whereas the chemotherapy medication used for patients in the Japanese cohort is uncertain.

**Figure 1:**
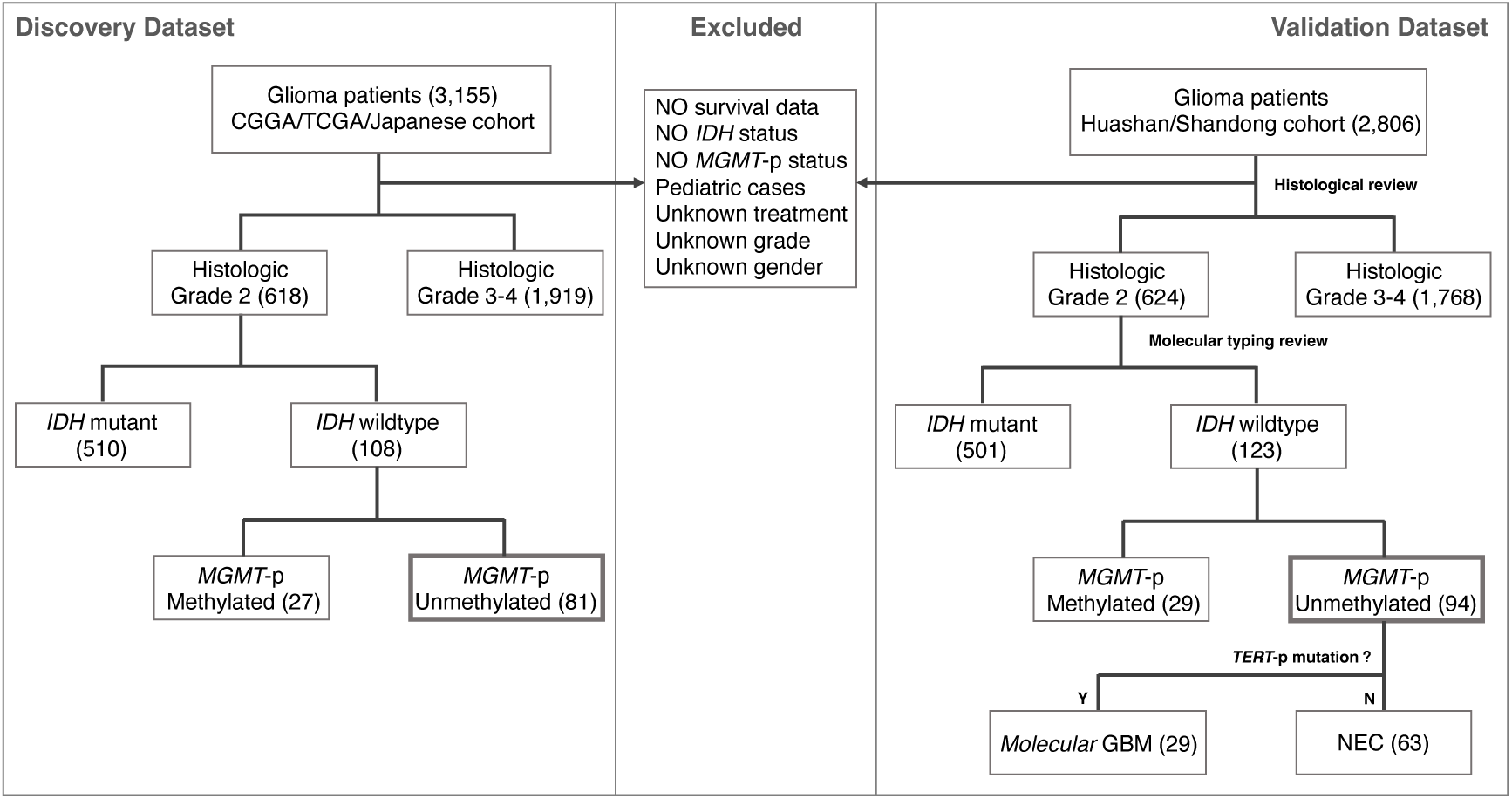
Sample inclusion and exclusion criteria. The Discovery dataset consisted of 108 patients with *IDH*wt histologic grade 2 gliomas from three public resources, including CGGA, TCGA and a Japanese cohort. The Validation dataset consisted of 123 patients with *IDH*wt grade 2 gliomas from two local institutions (Huashan Hospital and Shandong Provincial Hospital). Patients without available survival, molecular (*IDH* and *MGMT*-p) and required clinical (disease state, treatment and gender) information were excluded. CGGA, Chinese Glioma Genome Atlas; TCGA, The Cancer Genome Atlas; *IDH*, Isocitrate Dehydrogenase; *MGMT*-p, O^6^-methylguanine-DNA-methyltransferase promoter; WHO, World Health Organization

**Table 1.**
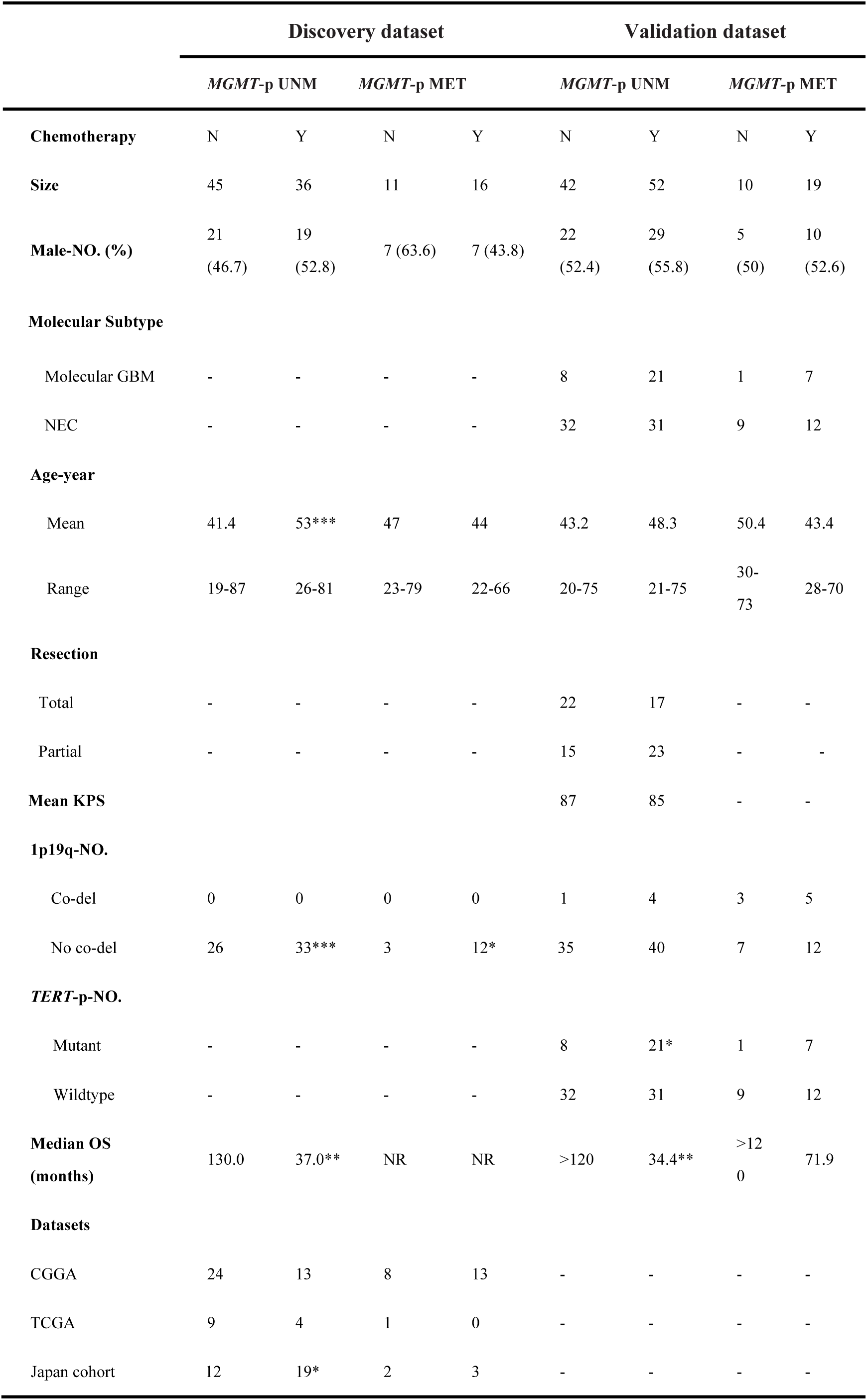
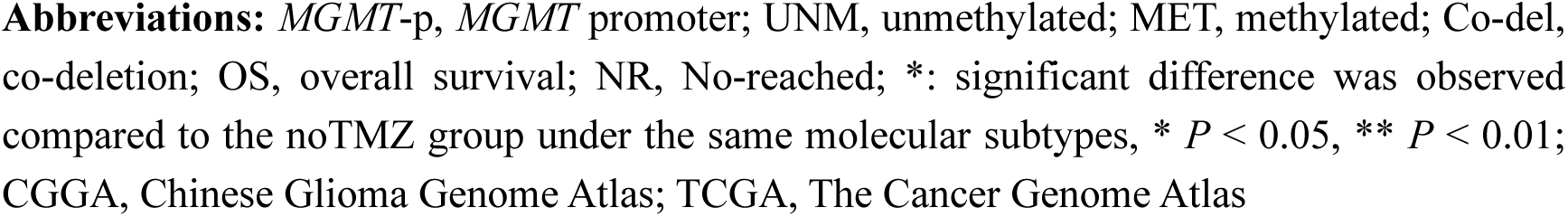
Clinical and molecular characteristics, and survival information for the Discovery and Validation datasets stratified by *MGMT*-p status when receiving or not receiving TMZ.

|  | Discovery dataset |  |  |  | Validation dataset |  |  |  |
| --- | --- | --- | --- | --- | --- | --- | --- | --- |
|  | *MGMT*-p UNM |  | *MGMT*-p MET |  | *MGMT*-p UNM |  | *MGMT*-p MET |  |
| **Chemotherapy** | N | Y | N | Y | N | Y | N | Y |
| **Size** | 45 | 36 | 11 | 16 | 42 | 52 | 10 | 19 |
| **Male-NO. (%)** | 21<br>(46.7) | 19<br>(52.8) | 7 (63.6) | 7 (43.8) | 22<br>(52.4) | 29<br>(55.8) | 5<br>(50) | 10<br>(52.6) |
| **Molecular Subtype** |  |  |  |  |  |  |  |  |
| Molecular GBM | - | - | - | - | 8 | 21 | 1 | 7 |
| NEC | - | - | - | - | 32 | 31 | 9 | 12 |
| **Age-year** |  |  |  |  |  |  |  |  |
| Mean | 41.4 | 53*** | 47 | 44 | 43.2 | 48.3 | 50.4 | 43.4 |
| Range | 19-87 | 26-81 | 23-79 | 22-66 | 20-75 | 21-75 | 30-73 | 28-70 |
| **Resection** |  |  |  |  |  |  |  |  |
| Total | - | - | - | - | 22 | 17 | - | - |
| Partial | - | - | - | - | 15 | 23 | - | - |
| **Mean KPS** |  |  |  |  | 87 | 85 | - | - |
| **1p19q-NO.** |  |  |  |  |  |  |  |  |
| Co-del | 0 | 0 | 0 | 0 | 1 | 4 | 3 | 5 |
| No co-del | 26 | 33*** | 3 | 12* | 35 | 40 | 7 | 12 |
| ***TERT*-p-NO.** |  |  |  |  |  |  |  |  |
| Mutant | - | - | - | - | 8 | 21* | 1 | 7 |
| Wildtype | - | - | - | - | 32 | 31 | 9 | 12 |
| **Median OS (months)** | 130.0 | 37.0** | NR | NR | >120 | 34.4** | >120 | 71.9 |
| **Datasets** |  |  |  |  |  |  |  |  |
| CGGA | 24 | 13 | 8 | 13 | - | - | - | - |
| TCGA | 9 | 4 | 1 | 0 | - | - | - | - |
| Japan cohort | 12 | 19* | 2 | 3 | - | - | - | - |

**Abbreviations:** *MGMT*-p, *MGMT* promoter; UNM, unmethylated; MET, methylated; Co-del, co-deletion; OS, overall survival; NR, No-reached; \*: significant difference was observed compared to the noTMZ group under the same molecular subtypes, \* $P < 0.05$ , \*\* $P < 0.01$ ; CGGA, Chinese Glioma Genome Atlas; TCGA, The Cancer Genome Atlas

The Validation dataset (n = 123, **Figure 1, Table 1**) was constructed to validate the survival differences with or without TMZ chemotherapy in patients with *IDH*wt histologic grade 2 gliomas. Patients diagnosed with grade 2 adult diffuse gliomas in 2010-2022 from Huashan Hospital of Fudan University and Shandong Provincial Hospital of Shandong First Medical University were included. Patients without available survival and required clinical information were excluded. Information on the maximum extent of resection (EOR), Karnofsky performance status (KPS), co-deletion of chromosomes 1p19q, and *TERT*-p mutation status was collected and used for adjusting the influence of interfering factors. MRI scans acquired at diagnosis were systematically reviewed to confirm the extent of resection. “NA” stands for missing patient data.

The study was approved by the Institutional Review Boards of Huashan Hospital (KY-2023-522) and the Biomedical Research Ethics Committee of Shandong Provincial Hospital (SWYX: NO. 2023-323). All tumor samples and clinical data were collected after written informed consent was obtained.

### TMZ treatment regimen

In the TMZ treatment group of the Validation dataset, 68 patients received the standard *Stupp* regimen, and three patients received TMZ chemotherapy alone. For both regimens, TMZ was administered orally at 200 mg/m(2)/day for five consecutive days and repeated every 28 days for six cycles. Further treatment was allowed at the discretion of the physicians for additional cycles. The noTMZ group consisted of 28 patients who received radiotherapy alone and 24 patients who were managed with a “wait-and-see” approach postoperatively.

### Molecular analysis

DNA was extracted from −80°C snap-frozen or formalin-fixed paraffin-embedded (FFPE) tumor samples. *IDH1* (R132H and R172H) and *IDH*2 mutation and *TERT* promoter mutation status were detected using a fluorescent PCR-capillary electrophoresis sequencing kit (SINOMD, Beijing, China). *MGMT* promoter methylation status was detected using a MSP-Fluorescent Probe Kit (SINOMD, Beijing, China).

### Molecular Subtyping

In the Validation dataset, patients were stratified into molecular GBM and NEC subtypes according to the WHO CNS5 guidelines to evaluate subtype-specific chemotherapy efficacy. Under these criteria, tumors harboring any of *TERT*-p mut, EGFR amp, or +7/−10 are classified as molecular GBM. However, due to sample limitations, complete information on EGFR and +7/−10 was not available. Evidence from prior molecular landscape studies indicates that EGFR amp and +7/−10 almost invariably co-occur with *TERT*-p mut, with fewer than 10% of cases presenting in isolation^2^. Moreover, *TERT*-p mut represents the predominant alteration among molecular GBMs^32^. On this basis, we adopted a pragmatic approach by classifying cases with *TERT*-p mut as molecular GBM and assigning all others to the NEC group. While this strategy may introduce a minor degree of misclassification, it is unlikely to affect the overall conclusions. Looking forward, we aim to address this limitation by collecting complete molecular information in prospective clinical studies.

### Omics Cohorts for Validation

We employed paired transcriptomic profiles from samples taken before and after recurrence, along with single-cell omics datasets, to validate the clinical observations. The paired cohort was prospectively collected at Huashan Hospital, consisting of patients with WHO grade 2 gliomas who all received postoperative TMZ and were followed for more than five years. Patients were stratified into recurrent and non-recurrent groups, and transcriptomic profiles were generated by next-generation sequencing. In addition, single-cell data were derived from patient samples previously collected at Huashan Hospital. Publicly available data from the CGGA325 dataset were also incorporated as an external validation cohort.

### Single-cell ssGSEA Analysis of MMR Pathway Activity

The gene set for the MMR pathway was obtained from the KEGG^33^ (https://www.kegg.jp) database. Single-cell RNA-sequencing (scRNA-seq) data were then analyzed to assess MMR pathway activity at the individual cell level. Specifically, the single-sample Gene Set Enrichment Analysis (ssGSEA)^34,35^ algorithm was applied to calculate an enrichment score for the MMR pathway for each cell. ssGSEA assigns a score based on the degree to which genes within the MMR pathway are coordinately upregulated or downregulated in each cell. The resulting MMR pathway scores were visualized using color gradients to reflect activity levels across different cells. Tumor cells were identified based on the expression of *SOX2* ^36^.

### Statistical methods

The survival analyses were performed using R packages *survival* (version 3.5.5) and *survminer* (version 0.4.9). The Log-rank test was used to quantify survival differences between groups. Categorical data were compared between subgroups using the Fisher’s exact test or the chi-square test. Comparisons for age, KPS and gene expression were performed using the Student’s t-test. Univariate and multivariate Cox regression analyses were used for risk assessment. A *P* value less than 0.05 was considered statistically significant. All statistical analyses were performed in R version 4.1.2.

## Results

### TMZ chemotherapy was associated with worse outcomes in *IDH*wt histologic grade 2 adult diffuse gliomas

Survival analyses were first performed to evaluate the effect of TMZ treatment on the survival of the overall cohort of patients with grade 2 *IDH*wt gliomas. Surprisingly, the OS of patients receiving TMZ was 84.1 months shorter than that of those without TMZ in the Discovery dataset (TMZ vs noTMZ, median OS: 45.9 months vs 130.0 months, Log-rank test *P* = 0.015, **Figure 2A**), whereas in the Validation dataset, the OS of patients receiving TMZ was 58.1 months shorter than that of those without TMZ (TMZ vs noTMZ, median OS: 71.9 months vs 130.0 months, *P* = 0.027, **Figure 2C**).

**Figure 2:**
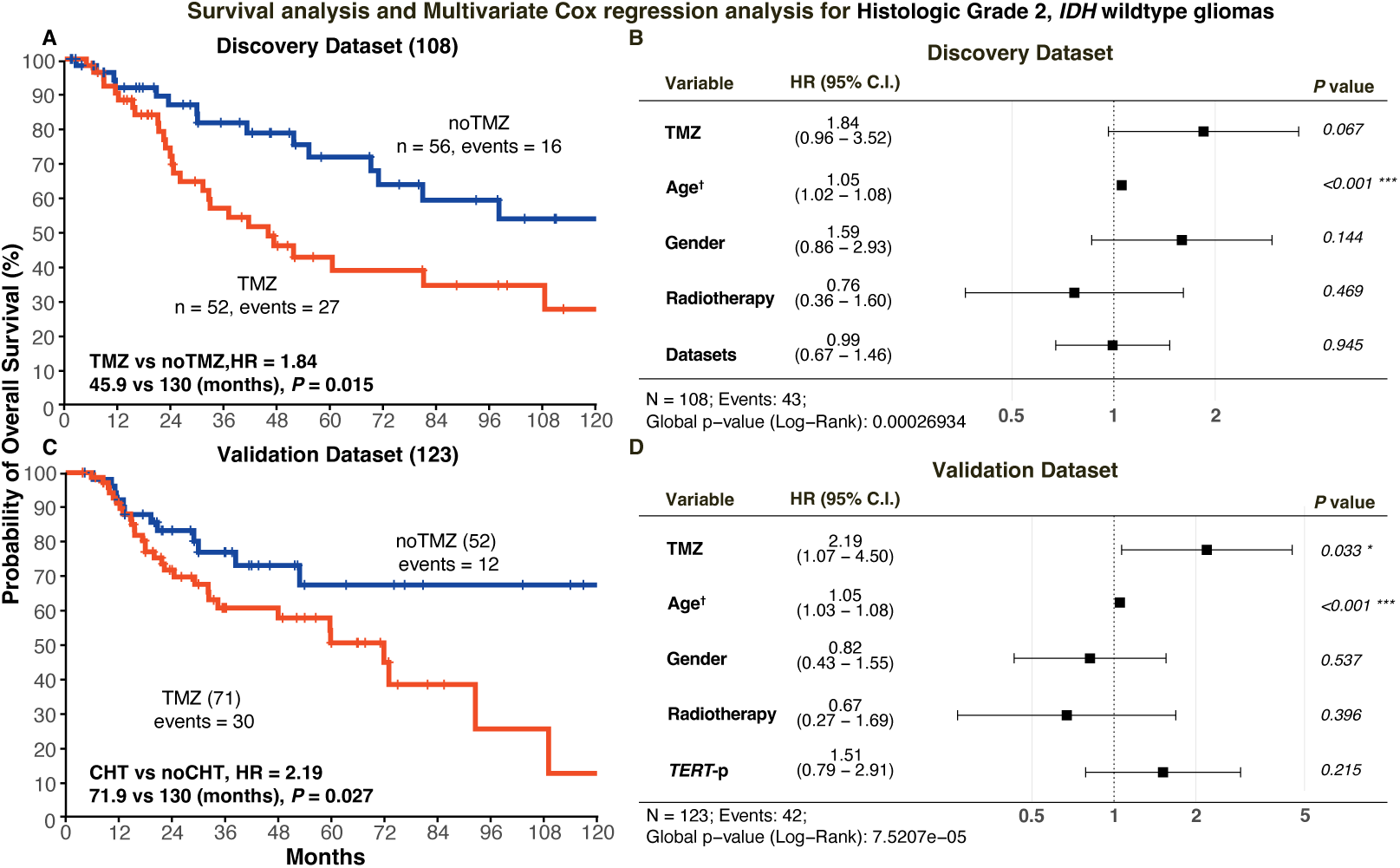
Kaplan-Meier analysis and multivariate cox regression analysis of overall survival (OS) stratified by whether receiving TMZ chemotherapy in patients with *IDH*wt grade 2 gliomas from the Discovery dataset and Validation dataset. A. Over five years of follow-up, the OS of patients receiving TMZ chemotherapy (TMZ, red line, n = 52) tended to be worse than those without TMZ (noTMZ, blue line, n = 56) (*P* = 0.015); B. Prognostic effects of TMZ together with other potential risk factors including age, gender, radiotherapy, and datasets were investigated in the Discovery dataset. TMZ was shown as a prognostic risk factor independent of other clinical factors in the *IDH*wt histologic grade 2 patients with marginally significance (*P* = 0.067). C. In the Validation dataset, The OS of patients receiving TMZ (red line, n = 71) showed significantly worse survival than those without TMZ (blue line, n = 52) (P = 0.027); D. Age, gender, Radiotherapy and *TERT*-p mutation status were added in the same analysis performed on the Validation dataset to correct for the effects of additional factors, and TMZ chemotherapy remained an independent high-risk factor (*P* = 0.033). Multi-Cox reference: noTMZ; Female; no-Radiotherapy; *TERT*-p wildtype; †Age: HR is for each one-year increase; *IDH*, Isocitrate Dehydrogenase; TMZ, receiving TMZ chemotherapy; noTMZ, not receiving chemotherapy; OS, overall survival

We compared baseline characteristics between patients who received TMZ and those who did not. In the Discovery dataset, significant differences were observed in age (TMZ vs noTMZ, n: 52 vs 56, 50.3 vs 42.4 years, *P* = 0.006) and in the proportion of patients without 1p/19q co-del (45/52 vs 29/56, *P* < 0.001), whereas such imbalances were not observed in the Validation cohort. However, in the Validation cohort, a significant imbalance was observed in the frequency of *TERT*-p mutations (28/71 vs 9/52, *P* = 0.015). To account for these baseline differences, we further performed multivariate Cox regression analyses. In the Discovery dataset, there was a marginally significant trend toward worse survival in patients receiving TMZ (HR = 1.84, 95%CI: 0.96–3.52, *P* = 0.067, Figures 2A, 2C). In the Validation dataset, TMZ therapy was again identified as a significant high-risk factor after adjusted for age, radiotherapy and Datasets (HR = 2.19, 95%CI: 1.07–4.50, *P* = 0.033, Figure 2B, 2D).

### TMZ correlated with worse survival only in *MGMT*-unmethylated gliomas

To evaluate the effect of *MGMT*-p methylation status on the survival of patients treated with TMZ, survival analyses were performed comparing patients with and without TMZ treatment within two molecular subgroups of *IDH*wt patients: *MGMT*-p unmethylated (WT_UNM, n = 81) and *MGMT*-p methylated (WT_MET, n = 27) (**Figure 3**). In the WT_UNM subgroup, patients receiving TMZ showed much worse survival than those without TMZ (Discovery dataset: TMZ vs noTMZ, 36 vs 45, median OS: 37.0 vs 130.0 months, Log-rank test *P* = 0.005; Validation dataset: TMZ vs noTMZ, 52 vs 42, median OS: 34.4 vs >120 months, *P* = 0.004, **Figure 3A, 3C**). In the WT_MET subgroups, no survival differences were observed between patients with and without TMZ treatment (Discovery dataset: TMZ vs noTMZ, 16 vs 11, median OS: >100 vs >120 months, *P* = 0.93; Validation dataset: TMZ vs noTMZ, 19 vs 10, median OS: 71.9 vs >120 months, *P* = 0.55, **Supplementary Figures 1A, 1B**).

**Figure 3:**
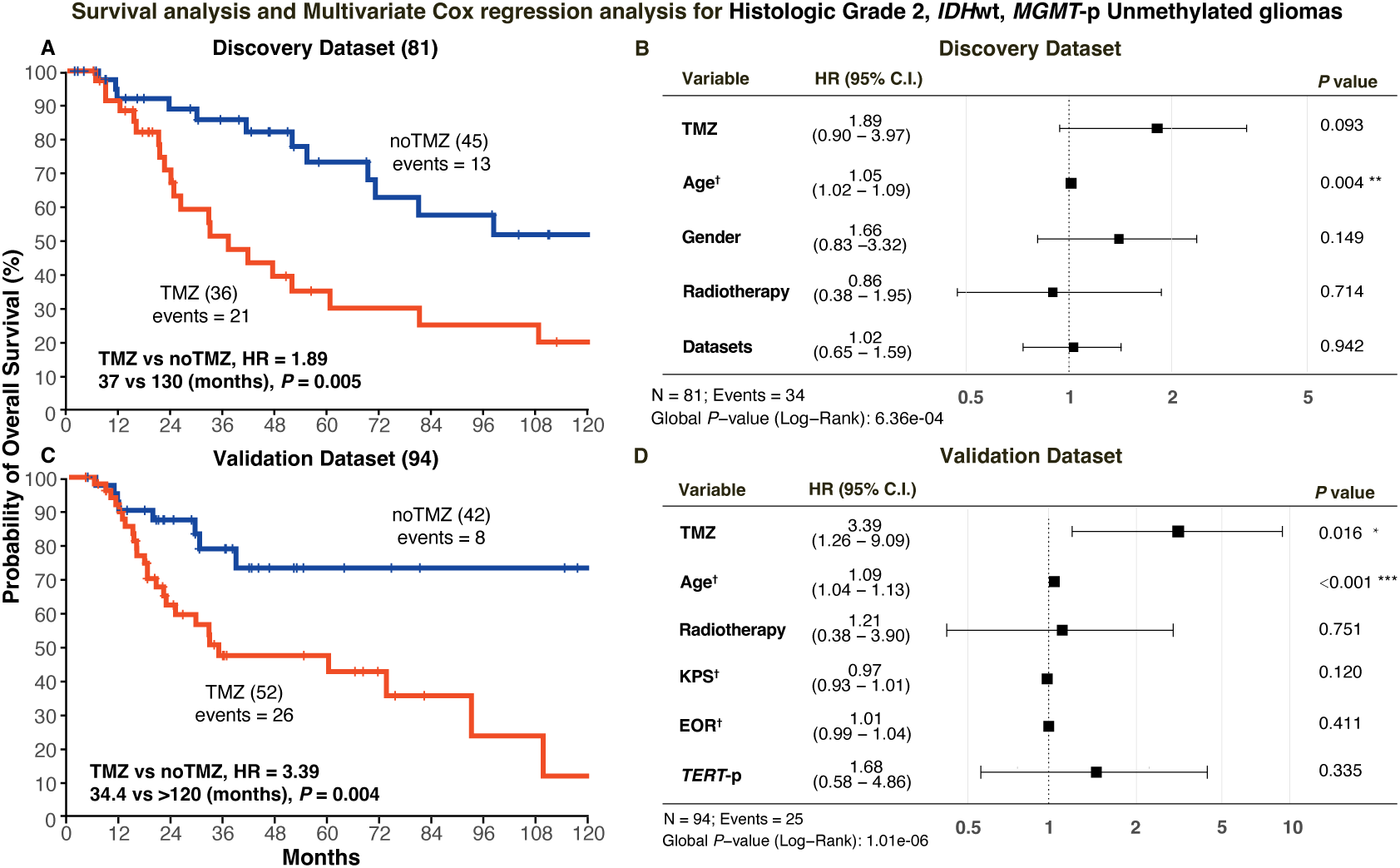
Kaplan-Meier analyses of OS stratified by whether patients received TMZ chemotherapy in patients with *IDH*wt grade 2 glioma and *MGMT*-p unmethylated, combined with multivariate Cox regression analysis in the Discovery and Validation datasets. A. A total of 81 patients with *IDH*wt and *MGMT*-p unmethylated grade 2 gliomas were included from the Discovery dataset. Kaplan-Meier analyses were conducted between those receiving TMZ or not. The OS of patients receiving TMZ (red line, n = 36) was significantly worse than that of those without TMZ (blue line, n = 45) in the WT_UNM subgroup of the Discovery dataset, *P* = 0.005. B. The prognostic effects of TMZ together with other potential risk factors including age, gender, radiotherapy, and datasets were investigated in the Discovery dataset. TMZ was an independent prognostic risk factor in the Discovery dataset. C. Kaplan-Meier analysis of overall survival of patients with WT_UNM in the Validation dataset (n = 94). The OS of patients receiving TMZ (red line, n = 52) was significantly worse than that of those without TMZ (blue line, n = 42), *P* = 0.004. D. *TERT*-p mutation status, EOR, and KPS were included in the multivariate analysis performed on the Validation dataset to adjust for the effects of additional factors, and TMZ chemotherapy remained an independent high-risk factor (*P* = 0.016). Multivariate Cox regression reference: no-Chemotherapy; Female; no-Radiotherapy; *TERT*-p wildtype; †Age: HR is for each one-year increase; †EOR: HR is for each 1% increase. †KPS: HR is for each 10-points increase; HR, hazard ratio; *IDH*, isocitrate dehydrogenase; *MGMT*-p, O6-methylguanine-DNA-methyltransferase promoter; wt, wildtype; TMZ, receiving TMZ chemotherapy; noTMZ, not receiving chemotherapy; OS, overall survival

Multivariate Cox regression analysis revealed TMZ therapy as an independent high-risk factor in the WT_UNM group. In the Discovery dataset, after adjustment for age, gender, and radiotherapy, TMZ therapy showed a borderline association with worse survival (HR = 1.89, *P* = 0.09, **Figure 3B**). In the Validation dataset, after adjustment for age, radiotherapy, *TERT*-p mutation status, EOR, and KPS, TMZ therapy remained an independent high-risk factor (HR = 3.39, *P* = 0.016, **Figure 3D**).

### “Molecular GBM” without *MGMT*-p methylation is not affected by TMZ chemotherapy

Since TMZ-associated risk was observed only in the WT_UNM subgroup, we further focused on gliomas with *MGMT*-p unmethylation. According to the WHO CNS5 guideline, we stratified patients in the Validation cohort into molecular GBM and NEC groups for further analysis: patients with *IDH*wt grade 2 gliomas harboring *TERT*-p mutation were categorized as molecular GBM, whereas those without *TERT*-p mutation were operationally classified as NEC. Survival analysis showed that patients receiving TMZ showed a trend toward worse survival in the molecular GBM subgroup, although no significant survival difference was observed (TMZ vs noTMZ, 8 vs 21, 20 vs 30 months, *P* = 0.37, Univariable Cox HR = 1.75, **Figure 4A**). In contrast, in the NEC group, TMZ remained significantly detrimental to survival (TMZ vs noTMZ, 32 vs 31, median OS 73 vs >120 months, *P* = 0.017, Univariable Cox HR = 3.56, **Figure 4B**)

**Figure 4:**
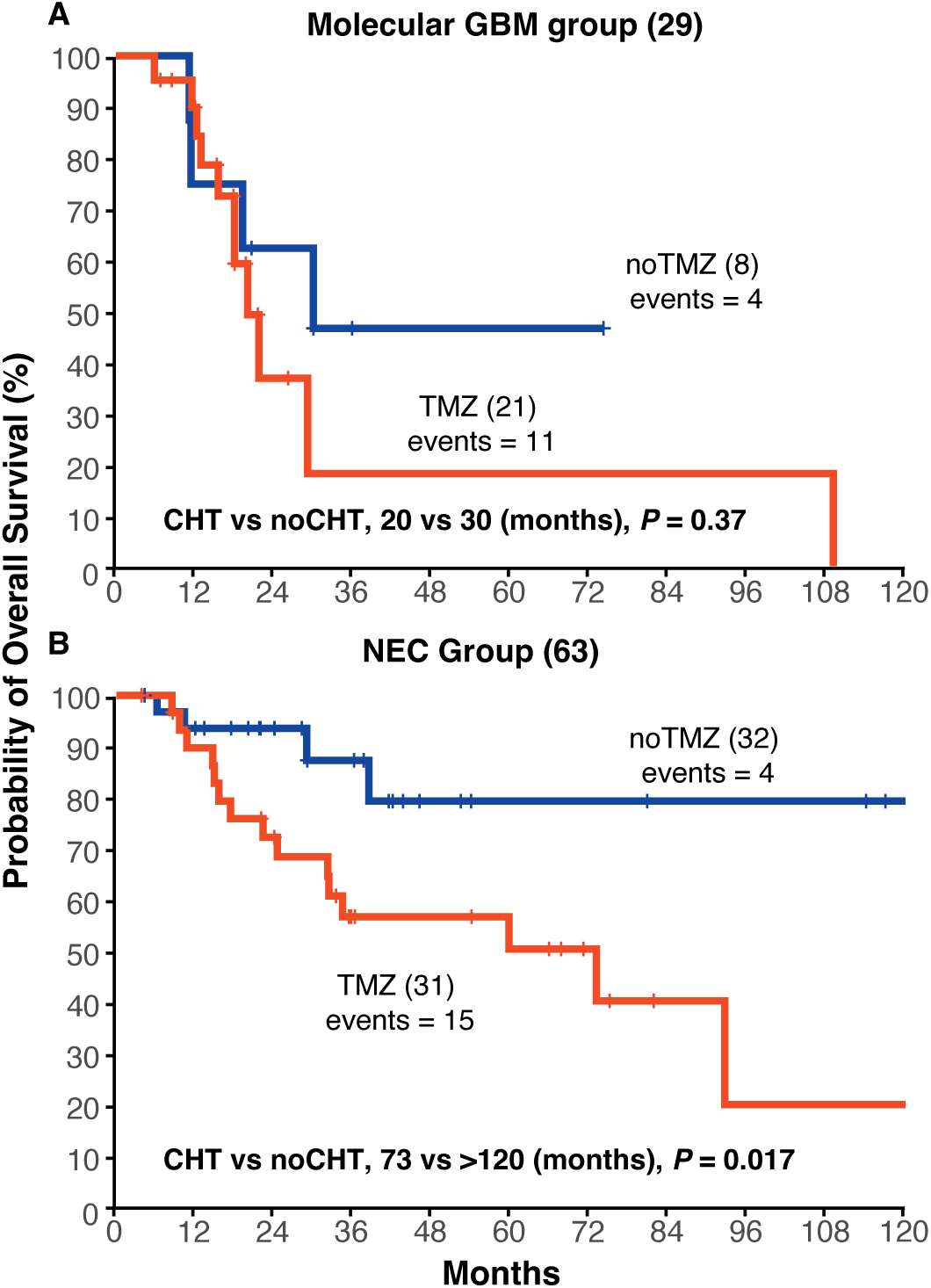
Kaplan-Meier analysis of overall survival (OS) stratified by the “molecular GBM group” and the “NEC Group ” in patients with *IDH*wt and *MGMT*-p unmet grade 2 gliomas from the Validation dataset. A: Kaplan-Meier analysis of overall survival of patients with “molecular GBM” in the Validation dataset, the OS of patients receiving TMZ (red line, n = 21) showed a worse prognostic trend than those without TMZ (blue line, n = 8), *P* = 0.37. B: Kaplan-Meier analysis was conducted comparing patients receiving TMZ therapy to those not receiving it in the NEC group. Patients receiving TMZ (red line, n = 31) had significantly worse OS than those not receiving TMZ (blue line, n = 32), *P* = 0.017. TMZ, receiving chemotherapy; noTMZ, not receiving chemotherapy; OS, overall survival

### Specific low MMR genes expression in *IDH*wt Grade 2 gliomas is associated with TMZ resistance

TMZ-induced O6-methylguanine exerts its cytotoxic effect through the combined action of *MGMT* and MMR, where reduced *MGMT* repair capacity and functional MMR signaling culminate in DNA double-strand breaks and cell death^23^. Lin et al.’s study summarized the combined impact of *MGMT* and MMR on TMZ resistance in gliomas, showing that cells are sensitive to TMZ chemotherapy only when they lack *MGMT* (*MGMT*-) and have normal MMR activity (MMR+). In gliomas, MMR activity is influenced by the expression of *MLH1*, *MSH2*, *MSH6*, and *PMS2* genes^37^. Therefore, we estimated MMR activity by analyzing the expression of MMR-related genes , which is critical for predicting TMZ sensitivity^23^.

Single-cell data indicate that in *IDH*wt grade 2 gliomas with unmethylated *MGMT*-p (TMZ-insensitive), *MGMT* is highly expressed in tumor cells, whereas MMR pathway activity is low. In contrast, in classic grade 4 GBM with methylated *MGMT*-p (TMZ-sensitive), *MGMT* expression is lower and MMR pathway activity is higher (**Figures 5A, 5B**). Analysis of a public dataset also revealed that *IDH*wt grade 2 gliomas display significantly lower expression of four key MMR-related genes compared to grade 3 gliomas and *IDH*mut grade 2 gliomas. This distinctive downregulation suggests a subgroup-specific reduction in MMR pathway activity, which may contribute to the limited efficacy of TMZ in these tumors (**Figure 5C**).

**Figure 5:**
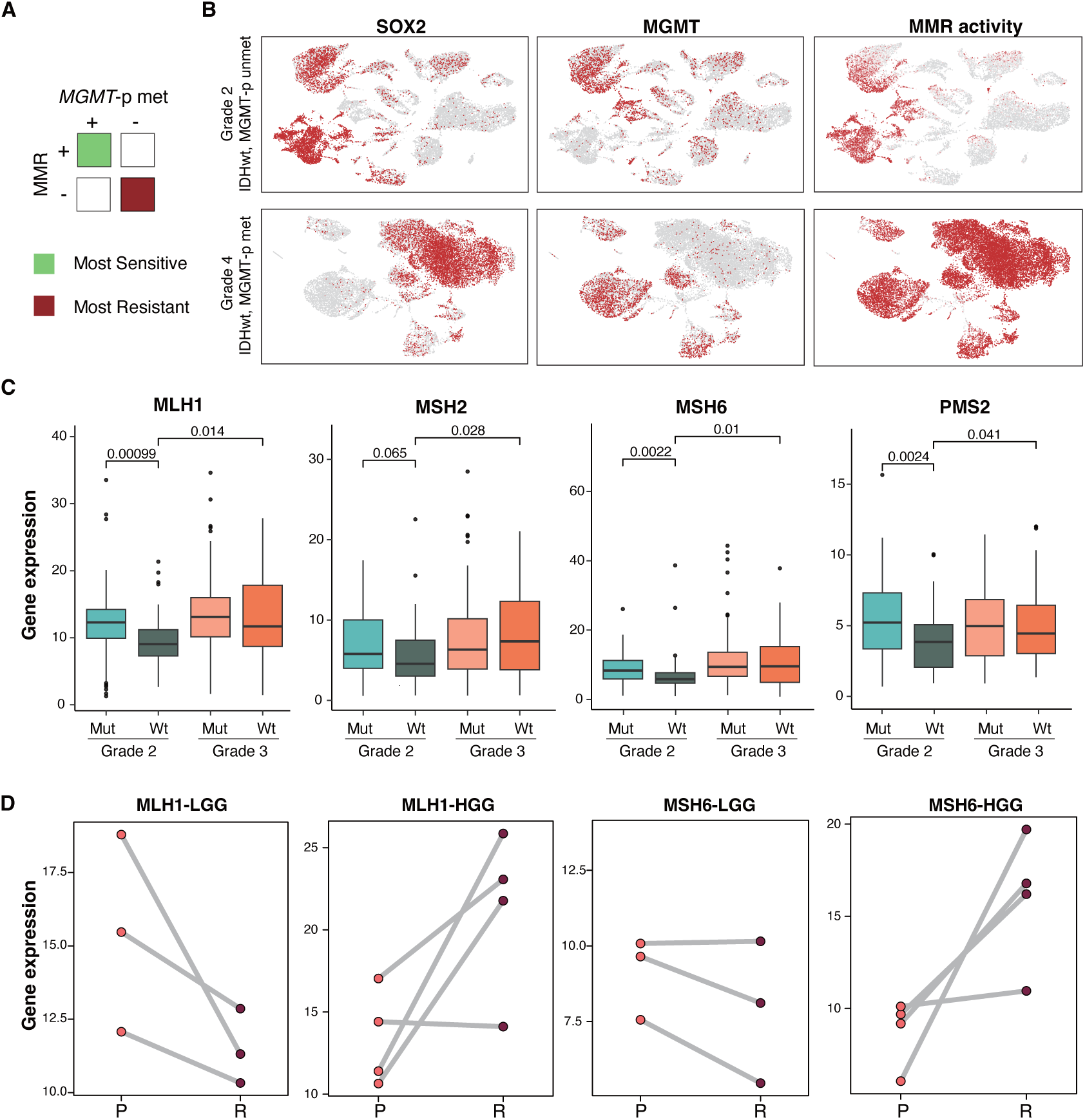
Specific low MMR pathway activity scores and MMR-related gene expression in *IDH*wt histologic grade 2 gliomas. Mismatch repair (MMR) pathway activity scores and MMR-related gene expression in histologic grade 2 glioma patients across different molecular subtypes are shown. A, B. Tumor regions of *IDH*wt grade 2 glioma patients exhibit low MMR pathway activity. Compared to classic grade 4 GBM, where tumor regions show low *MGMT* expression and high MMR pathway scores, *IDH*wt grade 2 gliomas display significantly lower MMR pathway scores. C. *IDH*wt grade 2 glioma patients exhibit the lowest expression of MMR-related genes compared to grade 3 gliomas and *IDH*-mutant grade 2 gliomas. D, E. Paired sample analysis shows that MMR gene expression remains stable upon recurrence as grade 2 or 3 (LGG), but increases significantly upon recurrence as grade 4 (HGG). MMR, mismatch repair; *MGMT*-p, O6-methylguanine-DNA-methyltransferase promoter; *IDH*, isocitrate dehydrogenase; P, Primary; R, Recurrent

Building on the observation of reduced MMR gene expression in *IDH*wt grade 2 gliomas, we further examined paired primary and post-recurrence samples from our local cohort. MMR gene expression decreased in low-grade recurrent patients (**Figure 5D**) but increased upon progression to grade 4 (**Figure 5E**). These findings suggest that TMZ provides limited benefit in low-grade gliomas, whereas its cytotoxic effect may become more pronounced as tumors progress to higher grades.

## Discussion

Due to the rarity of *IDH*wt grade 2 adult diffuse gliomas, previous studies could not draw definitive conclusions regarding the impact of TMZ on patient survival. In this study, we collected a large cohort of histologically confirmed *IDH*wt grade 2 gliomas and systematically evaluated the effect of postoperative TMZ chemotherapy. Our analysis revealed that early TMZ administration in patients with unmethylated *MGMT*-p was associated with reduced overall survival compared to patients without TMZ (median OS: 37 vs 130 months), even after adjusting for clinical factors such as EOR and KPS, suggesting potential adverse effects in this population. In contrast, TMZ treatment did not significantly affect OS in patients with grade 3/4 gliomas under the same stratification.

These findings highlight a subset of *IDH*wt grade 2 gliomas with favorable prognosis that may lack TMZ targets and experience detrimental effects from chemotherapy. From a prognostic perspective, *IDH*wt grade 2 patients who did not receive TMZ in our cohort showed markedly longer survival (>120 months) than previously reported^20^. Mechanistically, single-cell analyses indicated that in these low-grade tumors, *MGMT* is highly expressed in tumor cells while MMR pathway activity is low, in stark contrast to grade 4 GBM. Moreover, paired primary and post-recurrence samples showed that MMR gene expression increased only when tumors progressed to grade 4, suggesting limited TMZ benefit in low-grade disease and potential efficacy upon malignant progression.

According to WHO CNS5 guidelines and previous research, we stratified patients into molecular GBM and NEC subgroups mainly based on *TERT*-p mutation status^32^. In our retrospective cohort, we provide the first evidence that NEC patients with *MGMT*-p unmethylation are not suitable for immediate postoperative adjuvant TMZ, offering clinical guidance for this previously undefined subgroup. Notably, our data also indicate that histologic grade 2 molecular GBM with unmethylated *MGMT*-p derives no benefit from TMZ, as treated patients showed worse survival trends compared to untreated patients (TMZ vs noTMZ, 20 months vs. 30 months, *P* = 0.37).

TMZ exerts its cytotoxicity by generating O6-methylguanine adducts, which induce G/T mismatches in double-stranded DNA and trigger apoptosis when lesions accumulate^38^. We propose two main mechanisms underlying TMZ resistance in these lower-grade gliomas: (1) high *MGMT* expression promotes repair of alkylation damage, conferring drug resistance^39^, and (2) low MMR pathway activity limits recognition of DNA lesions, predisposing tumors to progression and recurrence. Although no MMR gene mutations were detected in our cohort, key MMR genes were downregulated compared to other subgroups. Paired pre- and post-recurrence analysis showed that MMR expression increased only upon progression to grade 4, indicating that TMZ re-administration may be more effective at later stages. Further studies are warranted to explore additional mechanisms driving MMR deficiency.

This study has several limitations. First, all cases in the Validation cohort were derived from a single institutional database, diagnosed between 2010 and 2022. During this period, clinical guidelines were updated three times, and physicians’ treatment decisions were not consistent, introducing potential selection bias; however, we can ascertain that their chemotherapy decisions were independent of *MGMT* methylation status and MMR activity. We attempted to mitigate this by incorporating and adjusting for clinical variables commonly influencing chemotherapy decisions, such as EOR and KPS. Nonetheless, prospective, multicenter, interventional studies with larger sample sizes are needed to validate our findings. Second, molecular GBM classification relied solely on *TERT*-p mutation status, while *EGFR* amplification and +7/–10 alterations were unavailable, potentially misclassifying a small subset (∼10%) of patients as NEC and introducing further bias. Comprehensive molecular profiling will be necessary in future studies to refine patient stratification.

In summary, our study demonstrates that combined high *MGMT* activity and MMR deficiency underlie the poor outcomes associated with temozolomide treatment in *IDH*-wildtype grade 2 gliomas. For the first time, in a large validation cohort, we show that patients with unmethylated *MGMT*-p derive no survival benefit from standard TMZ therapy and may even experience significantly worse outcomes. Notably, NEC patients within the unmethylated group exhibited markedly inferior survival when treated with chemotherapy, and molecular GBM with unmethylated *MGMT*-p also showed a trend toward poorer prognosis compared to untreated cases. Furthermore, we identified a characteristic downregulation of MMR genes in grade 2 *IDH*wt gliomas, which, together with high *MGMT* activity, likely contributes to TMZ resistance. Future studies should validate these findings in prospective, multicenter cohorts and explore alternative therapeutic strategies for this high-risk population.

## Disclosure

### Authors’ information

From the State Key Laboratory of Genetics and Development of Complex Phenotypes, Human Phenome Institute and School of Life Sciences, Shanghai Cancer Center (R.Z., Z.C., E.S., Q.C., S.S., Y.Z., L.S.), the Department of Neurosurgery, Huashan Hospital, Shanghai Medical College, Fudan University (S.W., C.L., J.W.), Shanghai Key Laboratory of Brain Function and Restoration and Neural Regeneration (S.W., C.L., J.W.), National Center for Neurological Disorders (S.W., C.L., J.W.); Neurosurgical Institute of Fudan University (S.W., C.L., J.W.) Department of Neurosurgery, Shandong Provincial Hospital Affiliated to Shandong First Medical University (S.S., Y.L., S.X.); Department of Neurosurgery, Affiliated Hospital of Southwest Medical University (L.C., Y.M.); Department of Bioinformatics, School of Medical Technology and Engineering, Key Laboratory of Medical Bioinformatics, Key Laboratory of Ministry of Education for Gastrointestinal Cancer, Fujian Medical University (W.Z.); and International Human Phenome Institutes (L.S.).

Dr. Jingsong Wu can be contacted at, Dr. Yingchao Liu at, Dr. Yuanting Zheng at.

### Authors’ contributions

J.W., Y.L., Y.Z., L.S., R.Z. and S.W. conceived the research and constructed the experimental design. J.W., Y.L., Y.Z. and L.S. managed the project. S.W., L.C., C.L., J.W., Y.L., Y.M. and S.X. provided the clinical information of local cohort. R.Z. and W.Z. completed the statistical analyses in the article. R.Z. and Z.C. drafted the initial version of the manuscript. E.S. and S.S. provided some critical ideas for data analysis and manuscript. L.S., Y.Z., E.S., W.Z., Z.C. and Q.C. contributed to the final revision of the paper. All authors have thoroughly reviewed and approved the final manuscript.

### Ethics approval and consent to participate

The study was approved by the Institutional Review Boards of Huashan Hospital (KY-2023-522) and the Biomedical Research Ethics Committee of Shandong Provincial Hospital (SWYX: NO.2023-323). All tumor samples and clinical data were collected upon written informed consent.

### Animal Studies

N/A.

### Availability of data and materials

Jinsong Wu had full access to all the data in the study and took responsibility for the integrity of the data and the accuracy of the data analysis. These data are available under a controlled access regimen to ensure the protection of personally identifiable data, and access can be obtained by contacting Prof. Jinsong Wu.

### Consent for publication

Not applicable

## Acknowledgements

Not applicable

## List of abbreviations

*EGFR*: Epidermal growth factor receptor
EOR: Maximum extent of resection
FFPE: Formalin-fixed paraffin-embedded
GBM: Glioblastoma
HR: Hazard Ratio
*IDH*: Isocitrate dehydrogenase
KPS: Karnofsky performance status
*MGMT*: O^6^-methylguanine-DNA methyltransferase
*MGMT*-p: O^6^-methylguanine-DNA methyltransferase promoter
MMR: Mismatch Repair
MRI: Magnetic resonance imaging
OS: Overall Survival
*TERT*: Telomerase reverse transcriptase
*TERT*-p: Telomerase reverse transcriptase promoter
TMZ: Temozolomide
WHO CNS5: 2021 WHO Classification of Tumors of the Central Nervous System5th edition
+7-10: The combination of gain of chromosome 7 and loss of chromosome 10

## Supplementary Figures

**Supplementary Figure 1:**
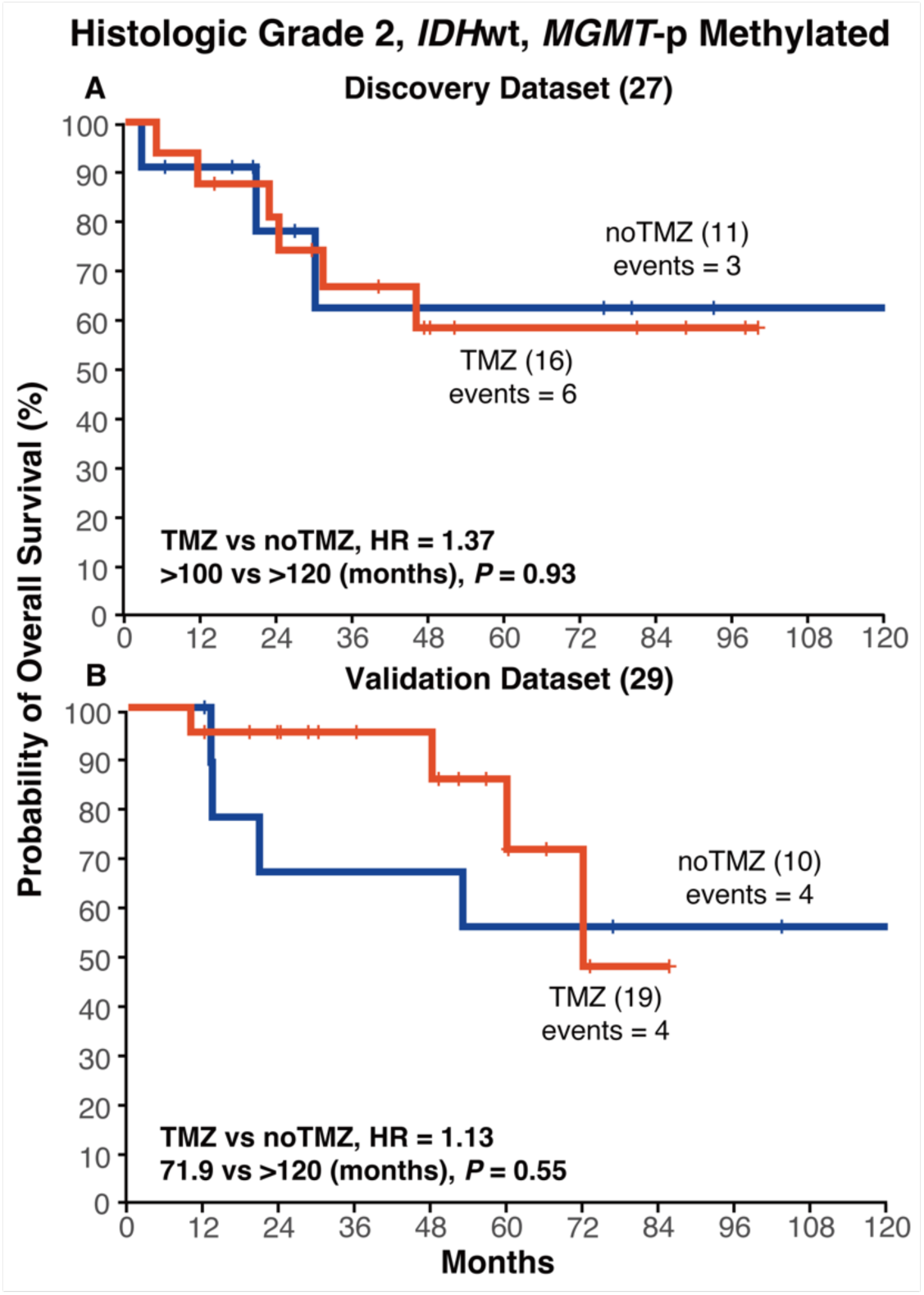
Kaplan-Meier analyses of OS stratified by whether receiving TMZ chemotherapy in patients with *IDH*wt and *MGMT*-p methylated grade 2 glioma. A: In the WT_MET of the Discovery dataset (N = 27), the OS did not differ between the TMZ and noTMZ groups, *P* = 0.93. B: In the WT_MET subgroup of the Validation dataset (N = 29), the OS of patients receiving TMZ did not differ from that of the noTMZ group, *P* = 0.55. *IDH*, isocitrate dehydrogenase; *MGMT*-p, O^6^-methylguanine-DNA-methyltransferase promoter; wt, wildtype; TMZ, receiving TMZ chemotherapy; noTMZ, not receiving chemotherapy; OS, overall survival

**Supplementary Figure 2:**
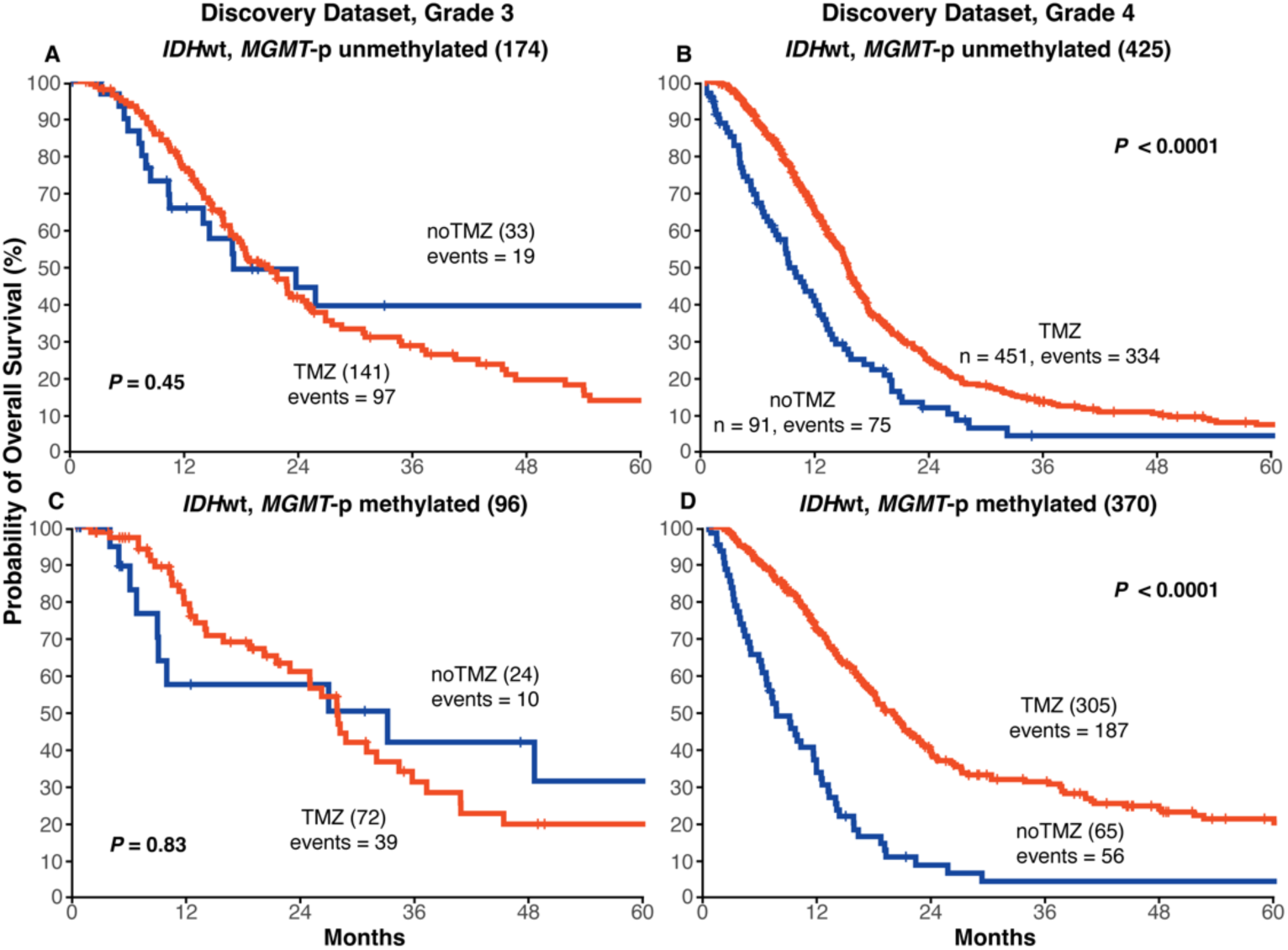
Different survival of patients with grade 3/4 gliomas stratified by *IDH*, *MGMT*-p and whether receiving TMZ. A-B: Kaplan-Meier analyses were conducted between those receiving TMZ therapy or not in histological grade 3 patients. The OS did not differ between TMZ and noTMZ groups in both “*IDH*wt, *MGMT*-p unmethylated” (A) and “*IDH*wt, *MGMT*-p methylated” (B). C-D: Kaplan-Meier analyses were conducted between those receiving TMZ therapy or not in histological grade 4 patients, the TMZ treatment patients had a significantly better prognosis in both “*IDH*wt, *MGMT*-p unmethylated” (C) and “*IDH*wt, *MGMT*-p methylated” (D) groups. IDH, isocitrate dehydrogenase; MGMT-p, O^6^-methylguanine-DNA-methyltransferase promoter; wt, wildtype; TMZ, receiving chemotherapy; noTMZ, not receiving chemotherapy; OS, overall survival

## Supplementary Table

**Table S1.** Review of Prospective and Retrospective Studies that TMZ chemotherapy in Grade 2 Gliomas.

| Article Number | Authors | Journal name | N | Response Rate | Age | M/F | High Risk | Seizures | Molecular | Median PFS | Year |
| --- | --- | --- | --- | --- | --- | --- | --- | --- | --- | --- | --- |
| 1 | Brada M, et al. | Ann Oncol.<sup>11</sup> | 30 | 10% | 40 | 13/17 | N | Y | - | - | 2003 |
| 2 | Quinn JA, et al. | J Clin Oncol.<sup>9</sup> | 46 | 61% | 41 | 27/19 | N | - | - | 22 | 2003 |
| 3 | Pace A, et al. | Ann Oncol.<sup>40</sup> | 43 | 47% | 40 | - | N | Y | - | 10 | 2003 |
| 4 | van den Bent MJ, et al. | J Clin Oncol.<sup>12</sup> | 39 | 52.6% | 49 | - | Y | - | - | 10.4 | 2003 |
| 5 | Hoang-Xuan K, et al. | J Clin Oncol.<sup>41</sup> | 60 | 17% | 43 | 36/24 | N | Y | 1p | - | 2004 |
| 6 | Levin N, et al. | Cancer.<sup>42</sup> | 28 | 35.7% | 38 | 17/11 | N | Y | 1p | - | 2006 |
| 7 | Everhard S, et al. | Ann Neurol.<sup>43</sup> | 68 | 7.4% | 41 | - | N | Y | MGMT-p | 28 | 2006 |
| 8 | Kaloshi G, et al. | Neurology.<sup>44</sup> | 149 | 15% | 44 | 84/65 | N | Y | 1p19q | 28 | 2007 |
| 9 | Dubbink HJ, et al. | Neurology.<sup>45</sup> | 49 | - | 38 | 29/20 | Y | - | - | 11 | 2009 |
| 10 | Kesari S, et al. | Clin Cancer Res.<sup>46</sup> | 44 | 20% | 43 | 30/14 | N | - | MGMT-p | 72 | 2009 |
| 11 | Houillier C, et al. | Neurology.<sup>47</sup> | 271 | 14.9% | 39 | 158/113 | N | - | IDH/1p19q | 34.6 | 2010 |
| 12 | Fisher BJ, et al. | Int J Radiat Oncol Biol Phys.<sup>8</sup> | 129 | - | 48 | 75/54 | Y | - | - | 54 | 2015 |
| 13 | Baumert BG, et al. | Lancet Oncol.<sup>10</sup> | 477 | - | 45 | 275/202 | Y | - | IDH | 39 | 2017 |
| 14 | Wahl M, et al. | Neuro Oncol.<sup>7</sup> | 120 | 6% | 39 | 67/53 | N | - | IDH/1p19q | 45.6 | 2017 |
| 15 | Bell EH, et al. | JAMA Oncol.<sup>21</sup> | 129 | - | 48 | 75/54 | N | - | MGMT-p | - | 2018 |
| 16 | Rudà R, et al. | J Neuro oncol. <sup>6</sup> | 60 | 38.3% | - | - | N | Y | MGMT-p | 40.8 | 2019 |
| 17 | Berzero G, et al. | Neuro Oncol. <sup>2</sup> | 9 | - | 55 | 36/11 | N | - | IDH | 59 | 2021 |

